# Relationship between body regional fat and cardiovascular disease in middle-aged and young adults: Results from NHANES 2011–2018 and Mendelian randomization meta-analysis

**DOI:** 10.1101/2024.03.19.24304562

**Authors:** Zheng Liu, Wenqi Shu, Meisi Liu, Wei Chong

## Abstract

**BACKGROUND:** To assess the correlation between regional body fat and cardiovascular diseases in middle-aged and young adults.

**METHODS:** Information on the regional body fat, including the mass and percentage of fat in the head, arms, trunk, and legs, was collected. Data for the observational study were derived from the National Health and Nutrition Examination Survey 2011–2018. The relationship between exposure and outcomes was primarily assessed using restricted cubic splines (RCS), weighted multivariable logistic regression, and subgroup analysis. In the Mendelian randomization (MR) analysis, the outcome was precisely defined as coronary heart disease (CHD), and the data primarily originated from the IEU OpenGWAS project.

**RESULTS:** A total of 10,158 participants aged 20–59 years were included, with a prevalence of cardiovascular diseases at 3.4%. Regional body fat was primarily associated with heart disease rather than with stroke. The RCS indicated a positive linear correlation between all body fat masses and the percentage of heart disease. After adjusting for confounding factors, logistic regression analysis revealed significant associations between heart disease and head fat mass (unit: 0.1 kg, odds ratio [OR] = 1.31, p = 0.002), head fat percentage (unit: 1%, OR = 1.37, p = 0.018), arm fat mass (unit: 1 kg, OR = 1.18, p = 0.047), and trunk fat mass (unit: 2.5 kg, OR = 1.14, p = 0.027). Leg fat mass and percentage of fat in the arms, trunk, and legs showed no significant correlation with heart disease (p > 0.05). The IEU OpenGWAS database does not include head-fat information. MR meta-analysis indicated that the fat mass and percentage in the arms, trunk, and legs were all causally related to CHD.

**CONCLUSION:** Head fat mass, head fat percentage, arm fat mass, and trunk fat mass were all associated with heart disease and were likely causally related.

**WHAT IS KNOWN:** Trunk fat mass is positively associated with certain cardiovascular risk factors, while leg fat mass is the opposite.

Dual-Energy X-ray Absorptiometry measures regional body fat, characterized by its speed, low radiation exposure, and cost-effectiveness, making it suitable for community settings.

**WHAT THE STUDY ADDS:** In middle-aged and young adults, regional body fat mass and percentage are not associated with stroke.

Head fat mass, head fat percentage, arm fat mass, and trunk fat mass are all associated with heart disease and are likely causally related.

Measurement of fat in the head, arms, and trunk may contribute to early screening for heart diseases in middle-aged and young adults.

## INTRODUCTION

Cardiovascular Disease (CVD) is a major cause of death and disease burden among residents in many countries worldwide, and the problem of premature death is becoming increasingly severe in many regions^1–4^. The United Nations member states hope to reduce the premature death rate of CVD by 25% by 2025^5^; however, it is difficult for all regions to achieve this goal^6^. Exploring the risk factors for CVD onset in middle-aged and young adults to facilitate early screening and timely intervention can help reduce the incidence and premature mortality in this population.

The distribution of fat in different regions is associated with various health risks. Early research found that trunk fat mass (TFM) and leg fat mass (LFM) are associated with certain risk factors for CVD^7–10^. These include metabolic syndrome^11,12^, insulin resistance^13,14^, hypertension^15^, and risk of systemic inflammation^16^, although the mechanisms are not yet fully understood. Most studies consider TFM to be a risk factor and LFM to be a protective factor, which has been confirmed in adolescents^17–19, 16,20^, and older adults^21,22^. However, these studies were observational and lacked causal inferences. There has been no research to confirm whether TFM and LFM truly influence the occurrence of CVD or to assess their potential as risk screening indicators for CVD, particularly in middle-aged and young adult populations. Body regional fat includes head fat mass (HFM) and arm fat mass (AFM); however, few studies have focused on HFM and AFM. Regional body fat was measured using Dual-Energy X-ray Absorptiometry (DXA). DXA is characterized by speed, ease of use, and low radiation exposure, making it suitable for primary care or physical examination settings.

The National Health and Nutrition Examination Survey (NHANES) is a cross-sectional survey based on a stratified population across the United States that provides a large-sample, nationally representative, and high-quality data. To date, no systematic study has been conducted on body regional fat mass (RFM) information in the NHANES 2011–2018. Mendelian randomization (MR) analysis is a tool that utilizes genetic data to analyze the causal relationship between exposure factors and outcomes^23^, with a lower risk of confounding bias and reverse causation^24^. Therefore, MR is considered a complementary method to randomized controlled trials, effectively serving as a strong supplement to observational studies.

The study design was depicted in **Figure 1**. In this study, we combined a large observational study from NHANES from 2011–2018 with an MR meta-analysis to comprehensively evaluate the relationship between body RFM (including HFM, AFM, TFM, and LFM) and CVD in middle-aged and young adult populations. Furthermore, we also analyzed the relationship between body regional fat percentage (RFP) and CVD.

**Figure 1.**
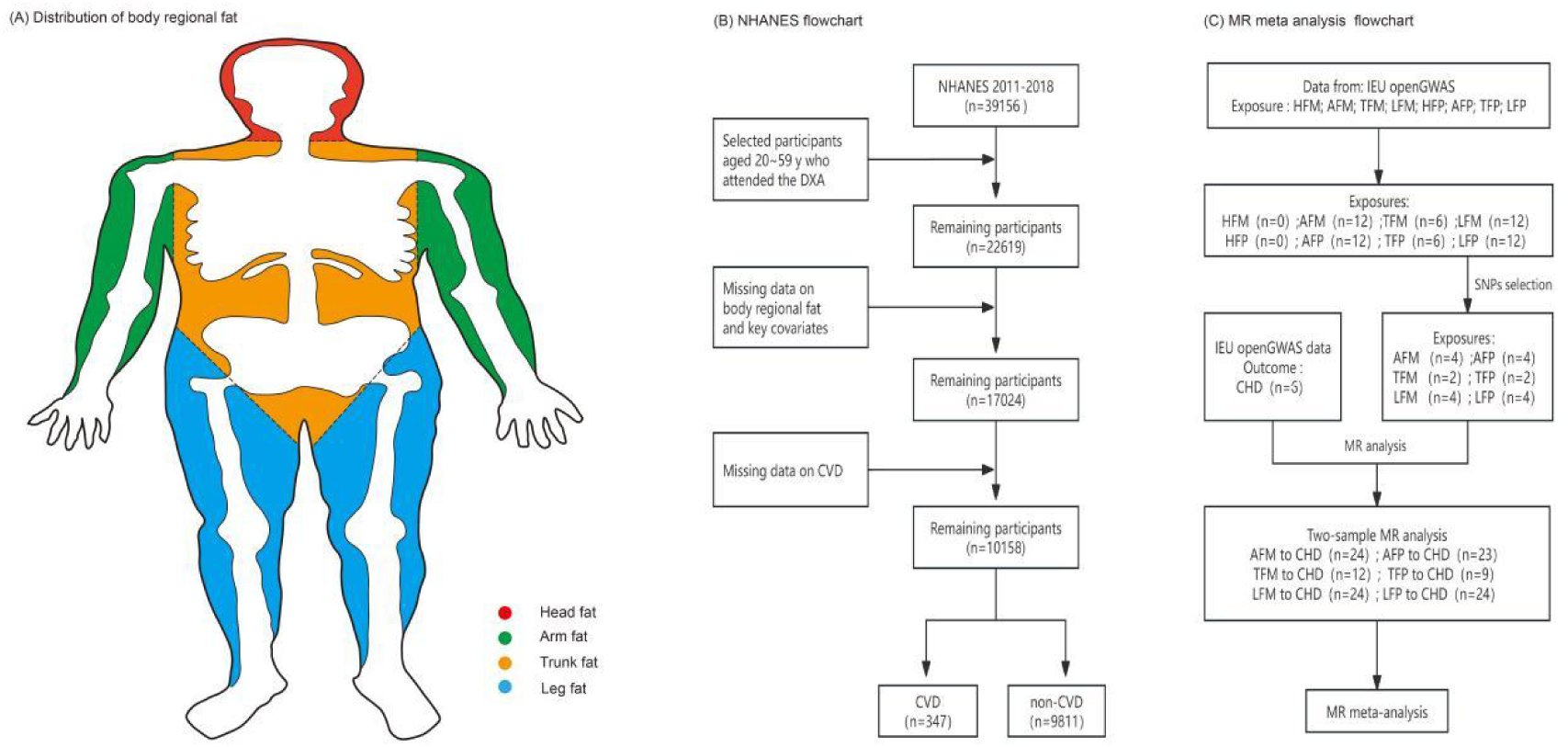
Schematic diagram of body region fat and study flowchart. NHANES, National Health and Nutrition Examination Survey; CVD, cardiovascular disease; CHD, coronary heart disease; DXA, Dual-Energy X-ray Absorptiometry; MR, mendelian randomization; SNPs, Single Nucleotide Polymorphisms; HFM, head fat mass; AFM, arm fat mass; TFM, trunk fat mass; LFM, leg fat mass; RFP, regional fat percentage; HFP, head fat percentage; AFP, arm fat percentage; TFP, trunk fat percentage; LFP, leg fat percentage.

## METHODS

### Cross-sectional study design

#### Study population

This study extracted population data from four cycles of the NHANES 2011-2018, as other cycles did not contain information on RFM and RFP. In the NHANES, the age distribution of the population undergoing RFM and RFP measurements ranged from 8 to 59 years. After excluding individuals with missing information on the main exposure factors and outcome indicators, the age distribution of the population was concentrated between 20 and 59 years old. Therefore, this study focused on the middle-aged and young adult populations. All participants in the NHANES provided informed consent, and the information was publicly anonymized and authorized by the NCHS Ethics Review Committee.

### Definition and extraction of body RFM, RFP, and CVD

The measurement of RFM and RFP was conducted using DXA. RFM included HFM, AFM, TFM, and LFM, with the original units in grams (g). RFP included head fat percentage (HFP), arm fat percentage (AFP), trunk fat percentage (TFP), and leg fat percentage (LFP). DXA measurements excluded individuals who were pregnant, weighed more than 450 pounds, or were taller than six feet five inches.

In this study, data on CVD were extracted from the Medical Conditions Questionnaire (MCQ) module in the NHANES, where MCQ information was obtained through personal interviews. In this study, CVD was defined to include congestive heart failure, coronary heart disease, angina, heart attack, and stroke, with the corresponding variables in MCQ being MCQ160B, MCQ160C, MCQ160D, MCQ160E, and MCQ160F. CVD includes other diseases as well, but owing to the lack of related information in the NHANES, CVD in this study was defined based on the aforementioned diseases.

### Definition of covariates

The main covariates in this study were age, sex, race, educational level, poverty level, smoking status, body mass index (BMI), obesity, diabetes, hypertension, and dyslipidemia. Smoking was defined as the smoking of at least 100 cigarettes per day. Obesity was defined as a BMI ≥ 28. Diabetes was defined based on a history of diabetes, hemoglobin A1c level ≥ 6.5%, or a fasting blood glucose level ≥ 126 mg/dL. Hypertension was defined based on a history of hypertension, the use of antihypertensive medication, or having a systolic blood pressure ≥ 140 mmHg or diastolic blood pressure ≥ 90 mmHg on at least 3 days in the examination module. Dyslipidemia included a history of lipid disorder, total cholesterol ≥ 6.2 mmol/L, low-density lipoprotein ≥ 1.4 mmol/L, high-density lipoprotein < 1.0 mmol/L, non-high-density lipoprotein ≥ 4.9 mmol/L, and triglycerides ≥ 2.3 mmol/L, with any one of these criteria being met.

### Statistical analysis

Continuous variables were presented as medians (IQR), while categorical variables were presented as n (%). The chi-square test with Rao & Scott’s second-order correction and the Wilcoxon rank-sum test were used for complex survey samples. Baseline investigations were conducted using CVD and RFM as separate outcomes. CVD was categorized as heart disease or stroke, and RFM was stratified into quartiles (Q1 to Q4). Pearson’s correlation coefficient analysis was conducted to examine the correlation between different RFMs in the body. Restricted cubic splines (RCS) were used to assess the linear relationship between exposure and outcome. Three logistic regression models were established to analyze the association between exposure and outcomes. The first model was unadjusted, the second model was further adjusted for age, sex, and race, and the third model was additionally adjusted for education, poverty level, BMI, and smoking. In the logistic regression analysis, the units of RFM were readjusted as follows: HFM to 0.1 kg, AFM to kg, TFM to 2.5 kg, and LFM to kg. Subgroup analyses were conducted based on sex, age, poverty level, smoking status, obesity, hypertension, diabetes, and dyslipidemia, and the interactions between exposure and subgroups were analyzed.

In this study, important variables with missing values were excluded, and no imputation was performed. All analyses of NHANES data were weighted to account for the complex sampling design. In this study, the minimum sample weight used was the MEC examination weight spanning four cycles. The weight variable was defined as 1/4WTMEC2YR. All statistical analyses were performed using R version 4.1.3 (The R Foundation for Statistical Computing, Vienna, Austria), with the significance level set at 0.05.

### MR meta-analysis

Based on the results of the cross-sectional study, exposure in the MR analysis was defined as RFM and RFP, and the outcome was defined as coronary heart disease (CHD), with data primarily sourced from the IEU OpenGWAS project. Initially, a two-sample MR analysis was conducted, in which the selected instrumental variables (Single Nucleotide Polymorphisms, SNPs) had to meet the following criteria: 1) genome-wide significance level (P < 5×10^-8); 2) not in linkage disequilibrium (r2 < 0.001, 10,000 kb); and 3) not a weak instrumental variable (F-statistic > 10). Five methods were used to determine the causal relationship between exposure and outcome: inverse variance-weighted (IVW), MR-Egger, weighted median, simple mode, and weighted mode. Among these, the IVW was the primary method, and secondary methods were used to evaluate the robustness of the IVW results. Heterogeneity was assessed using IVW and MR-Egger tests, and pleiotropy was evaluated using the MR-Egger intercept test. A sensitivity analysis was conducted using the leave-one-out method and funnel plots. The criteria for inclusion in the MR meta-analysis were that the odds ratio (OR) values from all five MR analysis methods were consistent in direction and that there was no horizontal pleiotropy. The MR meta-analysis was based on the IVW results. If there was heterogeneity in the statistical results, a random-effects model was used, and if there was no heterogeneity, a fixed-effects model was used.

## RESULTS

### NHANES population characteristics and baseline features

In total, 10,158 participants were included in this study, comprising 52% males and 48% females, with an average age of 39 years. In the weighted analysis, CVD accounted for 2.8%, heart disease for 2.1%, and stroke for 1.0%. As shown in **Table 1**, except for LFM and LFP, patients with CVD and heart disease had significantly higher HFM, AFM, TFM, HFP, AFP, and TFP values. Patients who experienced a stroke did not show differences in body RFM and RFP, with only AFP being slightly higher (24.07% vs. 24.29%, p = 0.003). Upon further dividing RFM into quartiles (Q1–Q4) for the outcome, it was observed that CVD and heart disease significantly increased with an increase in HFM, AFM, and TFM, while no difference was observed in LFM. Stroke patients with stroke showed a significant difference only in AFM (p = 0.022). This information can be found in **(TableS1-TableS4)**. AFM, TFM, and LFM exhibited very strong positive correlations, and HFM showed a positive correlation with these three variables; however, the correlation was relatively weaker **(Table 2)**.

**Table 1.**
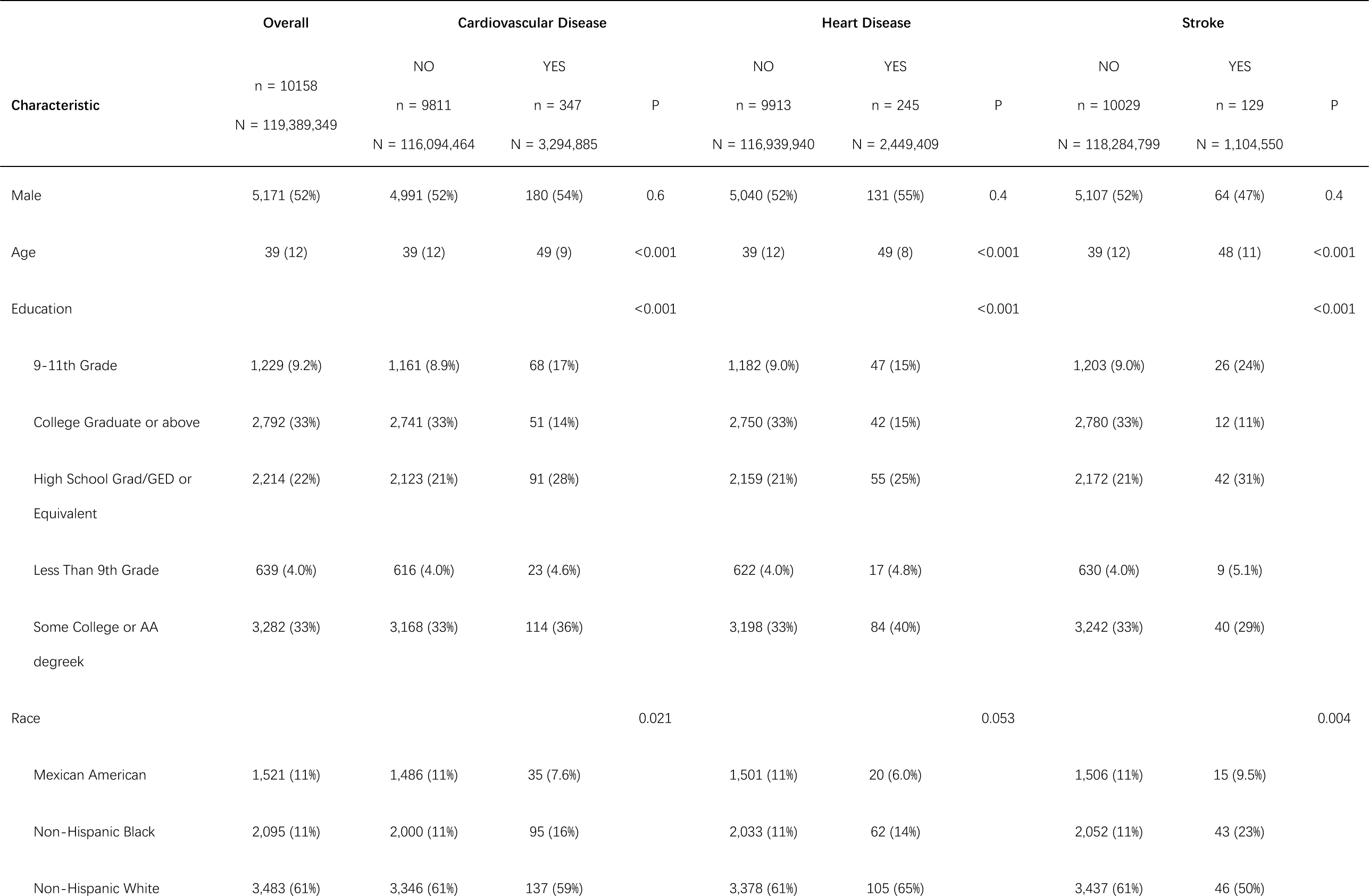
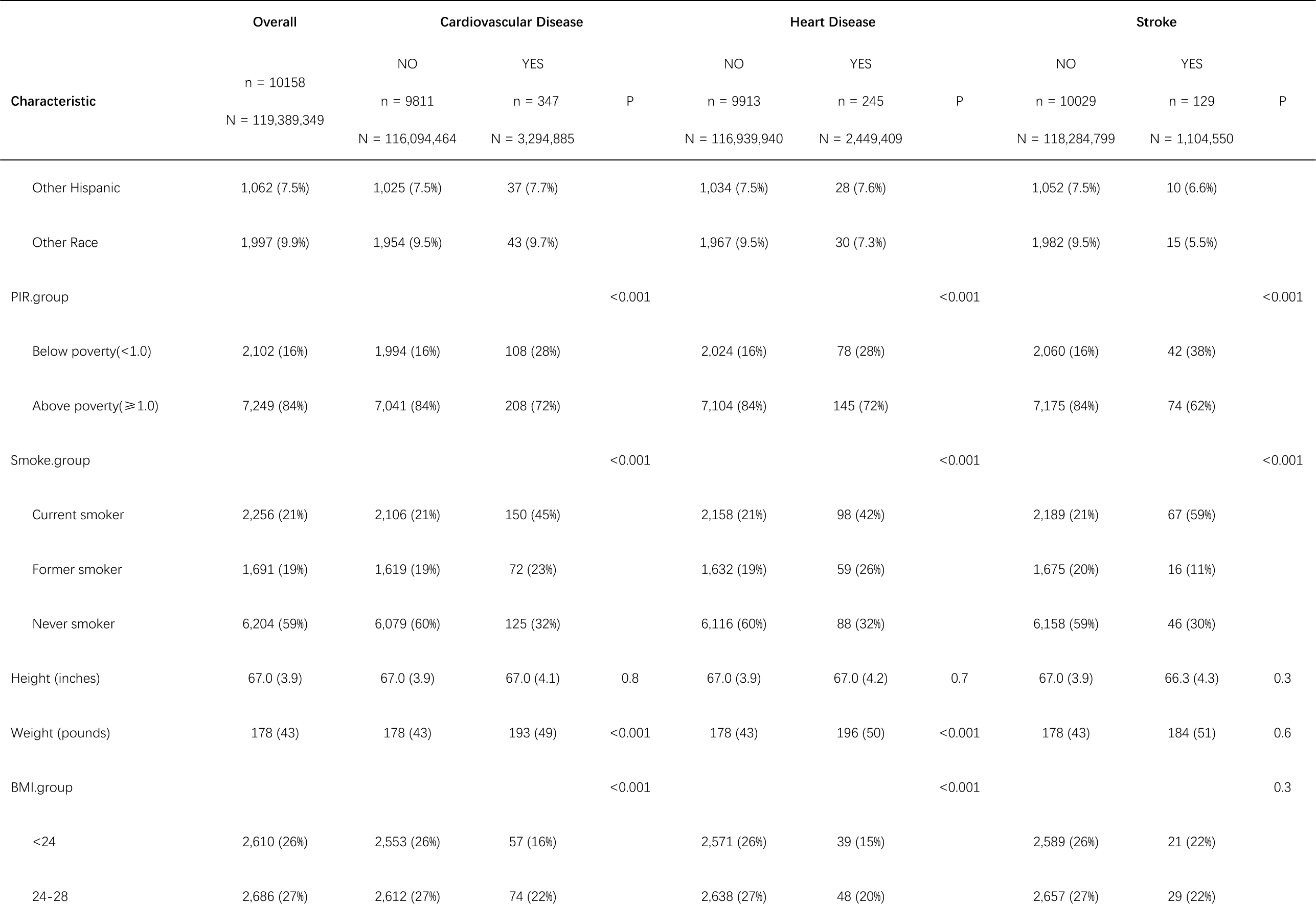
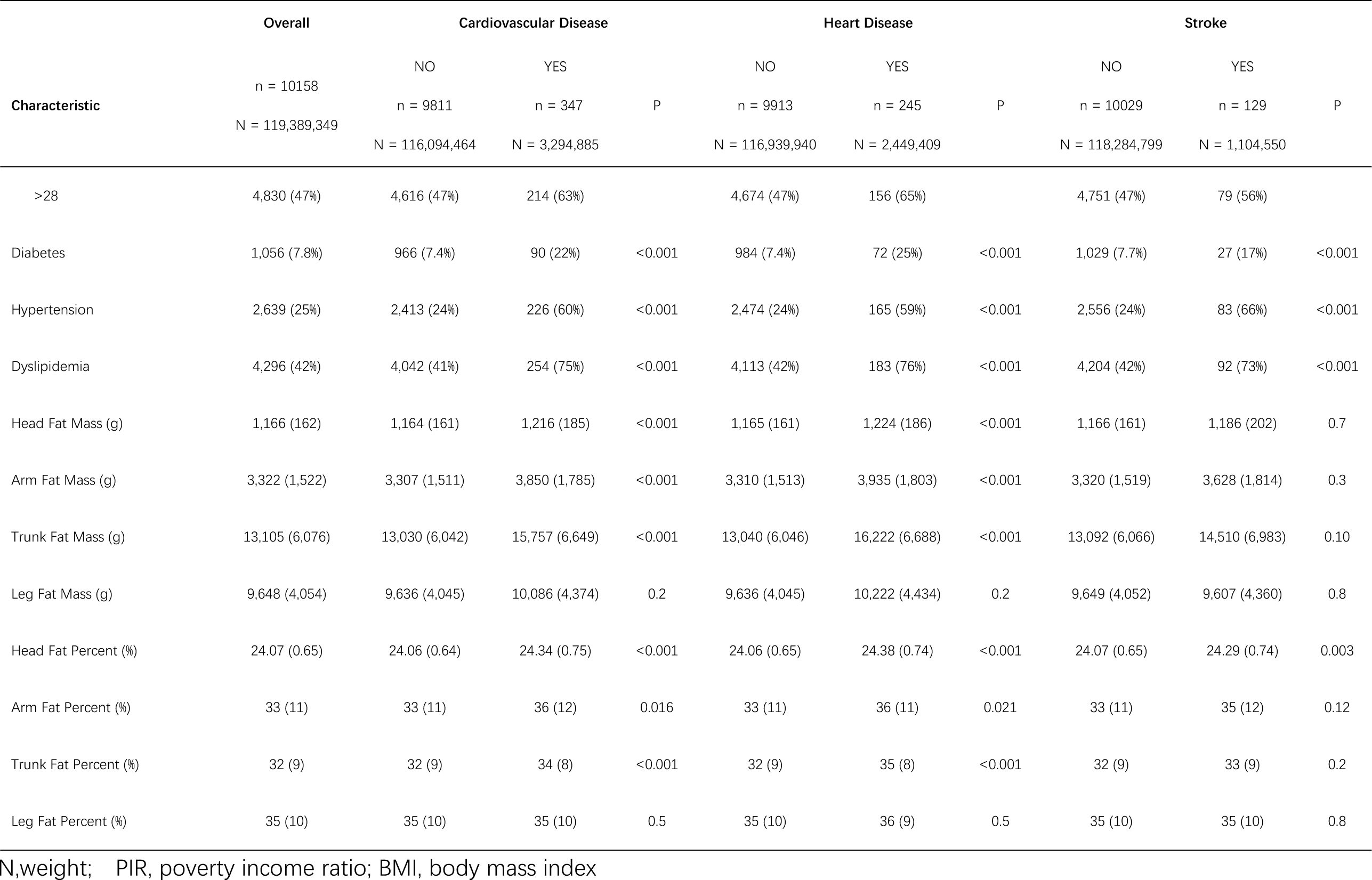
Baseline characteristics of the population.

**Table 2.** Correlation coefficients between fat mass in different body regions.

|  | Head Fat Mass | Arm Fat Mass | Trunk Fat Mass | Leg Fat Mass |
| --- | --- | --- | --- | --- |
| Head Fat Mass | 1 | 0.331975 | 0.426939 | 0.209801 |
| Arm Fat Mass | 0.331975 | 1 | 0.923785 | 0.856195 |
| Trunk Fat Mass | 0.426939 | 0.923785 | 1 | 0.796021 |
| Leg Fat Mass | 0.209801 | 0.856195 | 0.796021 | 1 |

### The relationship between RFM and RFP with heart disease in NHANES

As shown in **Figure 2**, the RCS indicated a positive linear correlation between heart disease and all four types of body RFM and RFP. **Table 3** shows the relationship between the four types of body RFM and RFP with heart disease under three models. Body RFM, HFM, AFM, and TFM showed consistent results across all three models. For every 0.1 kg increase in HFM, the risk of heart disease increased by 31% (OR = 1.31, 95% CI: 1.11–1.54, p = 0.002, Model 3). For every 1 kg increase in AFM, the risk of heart disease increased by 18% (OR = 1.18, 95% CI: 1.00–1.38, p = 0.047, Model 3). For every 2.5 kg increase in TFM, the risk of heart disease increased by 14% (OR = 1.14, 95% CI: 1.02–1.28, p = 0.027, Model 3). LFM was not significantly associated with heart disease in any of the three models. In terms of body RFP, only HFP showed consistent and significant results across all three models. For every 1% increase in HFP, the risk of heart disease increased by 37% (OR = 1.37, 95% CI: 1.06 –1.77, p = 0.018, Model 3). AFP and TFP were non-significant in Model 3, and LFP was not significant in any of the models.

**Figure 2.**
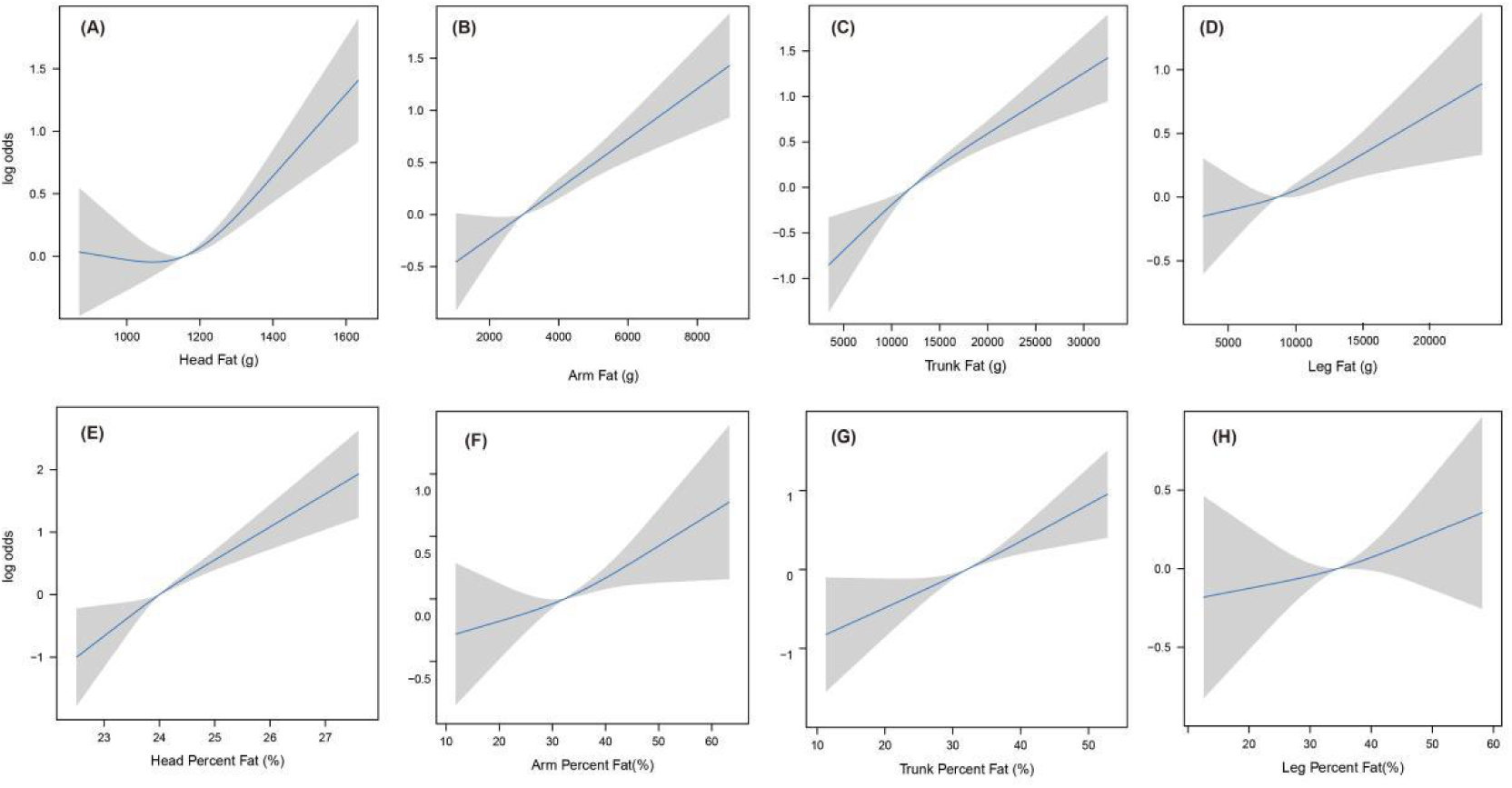
Restricted cubic spline analysis of the linear relationship between regional body fat and heart disease.

**Table 3.** Weighted logistic regression between fat mass and percentage in various body regions and heart disease.

|  | Model I OR (95%CI) | P | Model II OR (95%CI) | P | Model III OR (95%CI) | P |
| --- | --- | --- | --- | --- | --- | --- |
| **Regional Fat Mass** |  |  |  |  |  |  |
| Head Fat Mass (Unit:0.1kg) | 1.22 (1.09 , 1.37) | 0.001 | 1.31 (1.11 ,1.55) | 0.002 | 1.31 (1.11 ,1.54) | 0.002 |
| Arm Fat Mass (Unit: kg) | 1.24 (1.12 , 1.38) | <0.001 | 1.25 (1.10 ,1.41) | 0.001 | 1.18 (1.00 ,1.38) | 0.047 |
| Trunk Fat Mass (Unit:2.5kg) | 1.19 (1.12 , 1.28) | <0.001 | 1.17 (1.07 ,1.27) | <0.001 | 1.14 (1.02 ,1.28) | 0.027 |
| Leg Fat Mass (Unit: kg) | 1.03 (0.99 , 1.08) | 0.12 | 1.05 (1.00 , 1.11) | 0.069 | 1.01 (0.94 ,1.08) | 0.874 |
| **Regional Fat Percentage** |  |  |  |  |  |  |
| Head Fat Percent (%) | 1.68 (1.37 , 2.05) | <0.001 | 1.58 (1.23 , 2.03) | 0.001 | 1.37 (1.06 ,1.77) | 0.018 |
| Arm Fat Percent (%) | 1.02 (1.00 , 1.04) | 0.016 | 1.05 (1.01 , 1.08) | 0.009 | 1.03 (0.99 ,1.07) | 0.154 |
| Trunk Fat Percent (%) | 1.05 (1.02 , 1.07) | <0.001 | 1.05 (1.01 , 1.09) | 0.008 | 1.04 (0.99 ,1.09) | 0.138 |
| Leg Fat Percent (%) | 1.01 (0.99 , 1.02) | 0.428 | 1.02 (0.99 , 1.06) | 0.218 | 1.00 (0.96 ,1.04) | 0.801 |

Model I adjusted for: none
Model II adjusted for: age, gender, race
Model III adjusted for: age, gender, race, education, poverty income ratio, body mass index, smoking

The results of the subgroup analysis are shown in **Figure 3**. Interaction with the poverty income ratio (PIR) was present in all four types of RFM and RFP, hypertension interacted with AFM and TFM, and dyslipidemia interacted with AFM, TFM, LFM, TFP, and LFP. In the subgroup analyses, HFP showed the most consistent performance, whereas the associations of the rest of the RFM and RFP with heart disease weakened in some subgroups.

**Figure 3.**
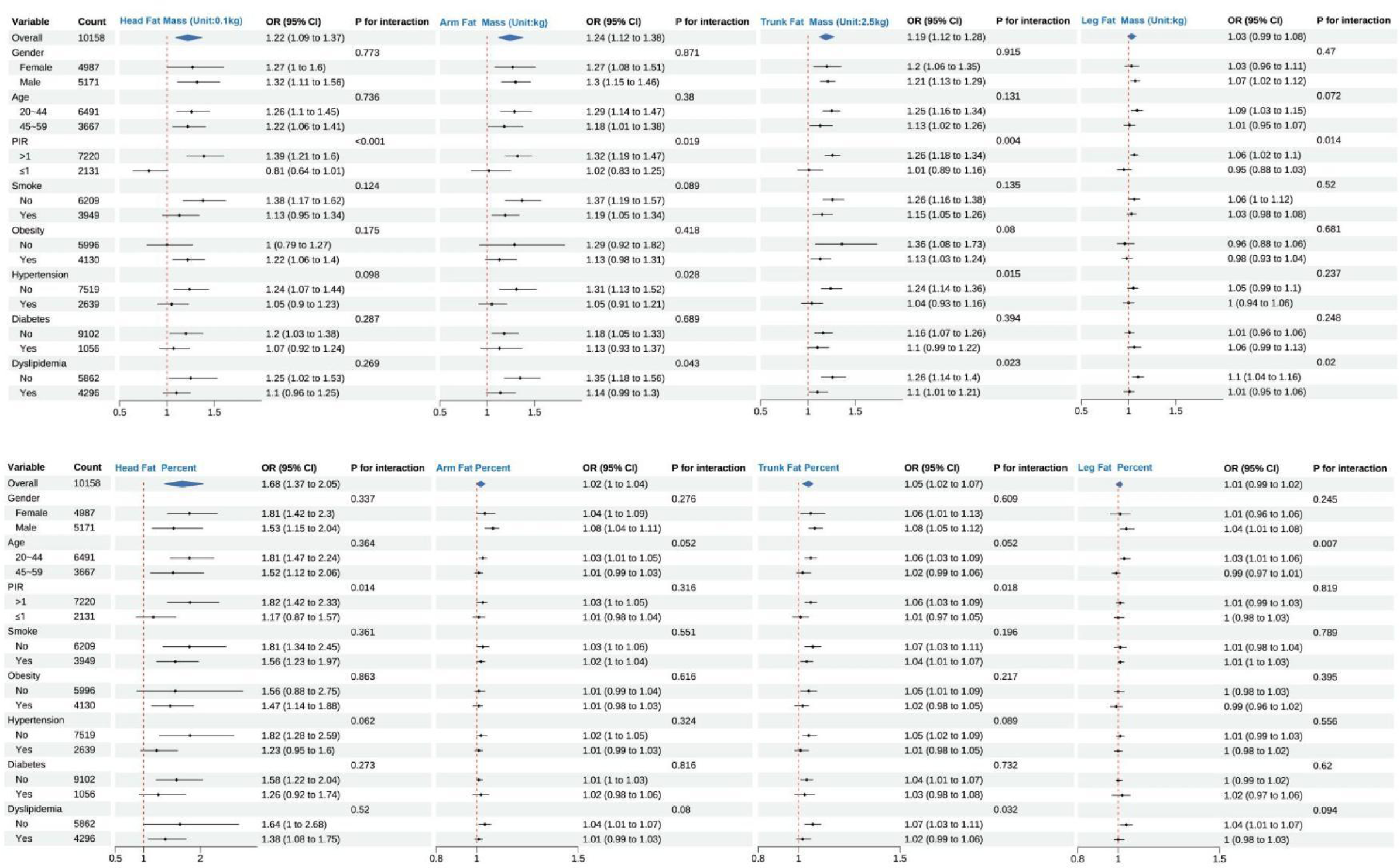
Subgroup analysis of the association between regional body fat and heart disease. PIR, poverty income ratio; OR, odds ratio.

### MR meta-analysis

No studies on HFM or HFP have been published in the IEU OpenGWAS databases. For AFM, LFM, AFP, and LFP, 12 studies each were identified, and for TFM and TFP, six studies each were found. After screening the SNPs, four studies on AFM, LFM, AFP, and LFP and two studies on TFM and TFP met the criteria. Based on the results of the observational study, we chose CHD as the exposure source and identified six studies in the IEU OpenGWAS database. The results of each MR analysis are detailed in the **TableS5 to TableS10**. Heterogeneity was present in all MR meta-analyses in this study (p < 0.05); hence, all were based on random effects models. The MR meta-analysis results showed a causal relationship between AFM and CHD (OR = 1.174, 95% CI: 1.092–1.261, p < 0.001), between TFM and CHD (OR = 1.084, 95% CI: 1.022–1.149, p = 0.007), between LFM and CHD (OR = 1.161, 95% CI: 1.083–1.245, p < 0.001), between AFP and CHD (OR = 1.260, 95% CI: 1.139–1.134, p < 0.001), between TFP and CHD (OR = 1.170, 95% CI: 1.044–1.310, p = 0.007), and between LFP and CHD (OR = 1.237, 95% CI: 1.119–1.367, p < 0.001).

## DISCUSSION

This study explored, for the first time, the relationship between regional body fat and CVD in the middle-aged and young populations using data from the NHANES 2011–2018 and MR meta-analysis. An observational study showed that HFM, AFM, TFM, and HFP were significantly associated with heart disease, whereas AFP and TFP had a weaker association, and LFM and LFP had no association with heart disease. An observational study also found that regional body fat was not associated with stroke. As the primary heart disease in the observational study was CHD, the MR meta-analysis selected CHD as the outcome and confirmed the causal relationship between AFM, TFM, AFP, and TFP and CHD. However, the MR meta-analysis results for LFM and LFP were inconsistent with those of observational studies. Owing to a lack of data, MR analyses of HFM and HFP were not conducted. The findings of this study are important for early screening and prevention of heart disease in middle-aged and young individuals. The detection of body regional fat is characterized by low cost, low radiation exposure, convenience, and speed, making it suitable for screening in the community. By focusing on fat accumulation in specific body regions, doctors may be able to identify high-risk individuals more accurately and implement targeted intervention measures.

Regional body fat refers to the fat accumulated in specific areas of the body. It can accurately differentiate between fat and nonfat tissues (such as muscles), providing detailed information about fat distribution. The distribution of fat in different regions is associated with various health risks; hence, regional body fat offers a more comprehensive health assessment. Previous studies have discovered associations between TFM, LFM, and certain cardiovascular health factors, which inspired the scientific hypothesis for this study. Notably, the previously understudied HFM and HFP levels were identified as significant risk factors for heart disease. Based on the results of the baseline analysis, stroke, whether as an outcome or a contributing factor, showed no significant correlation with fat in any of the four body regions **(Table 1, Table S1-Table S4)**. Therefore, in subsequent studies, we specifically refined the outcomes of CVD to include heart disease. This study aimed to determine the strength of the causal relationship between regional body fat and heart disease to aid in early screening of heart disease. We did not intend to prove this to be an independent risk factor. Therefore, excessive confounding factors such as hypertension, dyslipidemia, and diabetes were not adjusted for. Instead, a subgroup analysis was used as a substitute. Next, we discuss the results of head fat, arm fat, trunk fat, and leg fat in this study in sequence.

Previous studies, which are few in number on head fat, have found that the accumulation of head fat is closely related to dyslipidemia and hyperuricemia^25^, consistent with the results of this study. The baseline survey in this study showed that an increase in HFM was not only associated with heart disease, but also significantly correlated with hypertension, dyslipidemia, and diabetes **(TableS1).** While there is currently limited research on head fat, we believe that it affects the occurrence of CHD by influencing certain cardiovascular risk factors, similar to trunk fat. In this study, after adjustment, HFM and HFP were not only the most stable among all indicators but were also the most clinically significant **(Table3)**. In all subgroup analyses of the PIR, there was evidence of an interaction, which was an interesting finding. This suggests that differences in socioeconomic status may lead to variations in various lifestyles and health behaviors, potentially influencing the risk of heart disease. The association between HFM and heart disease weakened in some subgroups, possibly because other risk factors present in these subgroups diminished the effect of HFM. However, HFP showed stable performance across all subgroups. Unfortunately, there have been no studies on HFM and HFP in GWAS databases. However, by combining observational studies with MR meta-analyses of other regional fats, we believe that head fat, especially HFP, has great potential as a screening indicator for heart disease risk.

Although both are limb fats, compared to leg fat, there are fewer studies on arm fat and its association with health risks, and the findings are contradictory. Sánchez et al.’s cross-sectional study of 683 university students aged 18–30 found no association between arm fat and risk factors such as dyslipidemia, insulin resistance, and hypertension^26^. In a study of 9,930 middle-aged and older individuals, Shi et al. found that mid-upper arm circumference was positively correlated with metabolic syndrome^27^. Gavi et al. found a negative correlation between the limb fat-to-trunk fat ratio (LFM/TFM) and insulin resistance risk in older adults ^28^, whereas Travis et al. found no association between LFM/TFM and any markers of cardiovascular metabolic risk in older individuals^21^. However, it is important to note that both of these studies had relatively small sample sizes, with 38 participants in the former and 136 in the latter, and did not differentiate between arm and leg fat but instead studied limb fat collectively. Leg fat is traditionally recognized as a protective factor for health and combining arm fat and leg fat with limb fat is inappropriate. The baseline survey of this study revealed a significant association between the occurrence of heart disease and high AFM and AFP **(Table1)**. High AFM was significantly associated with heart disease and cardiovascular risk factors **(TableS2)**, as indicated by the RCS curves, which showed a clear positive linear correlation **(Figure2B, 2F)**. In further logistic regression and subgroup analyses, AFM performance remained stable, whereas AFP levels showed weaker statistical and clinical significance. MR meta-analysis further confirmed the causal relationship between AFM and CHD **(Figure 4B)**. Therefore, we believe that AFM is a risk factor for heart disease and has clinical value in risk screening.

**Figure 4.**
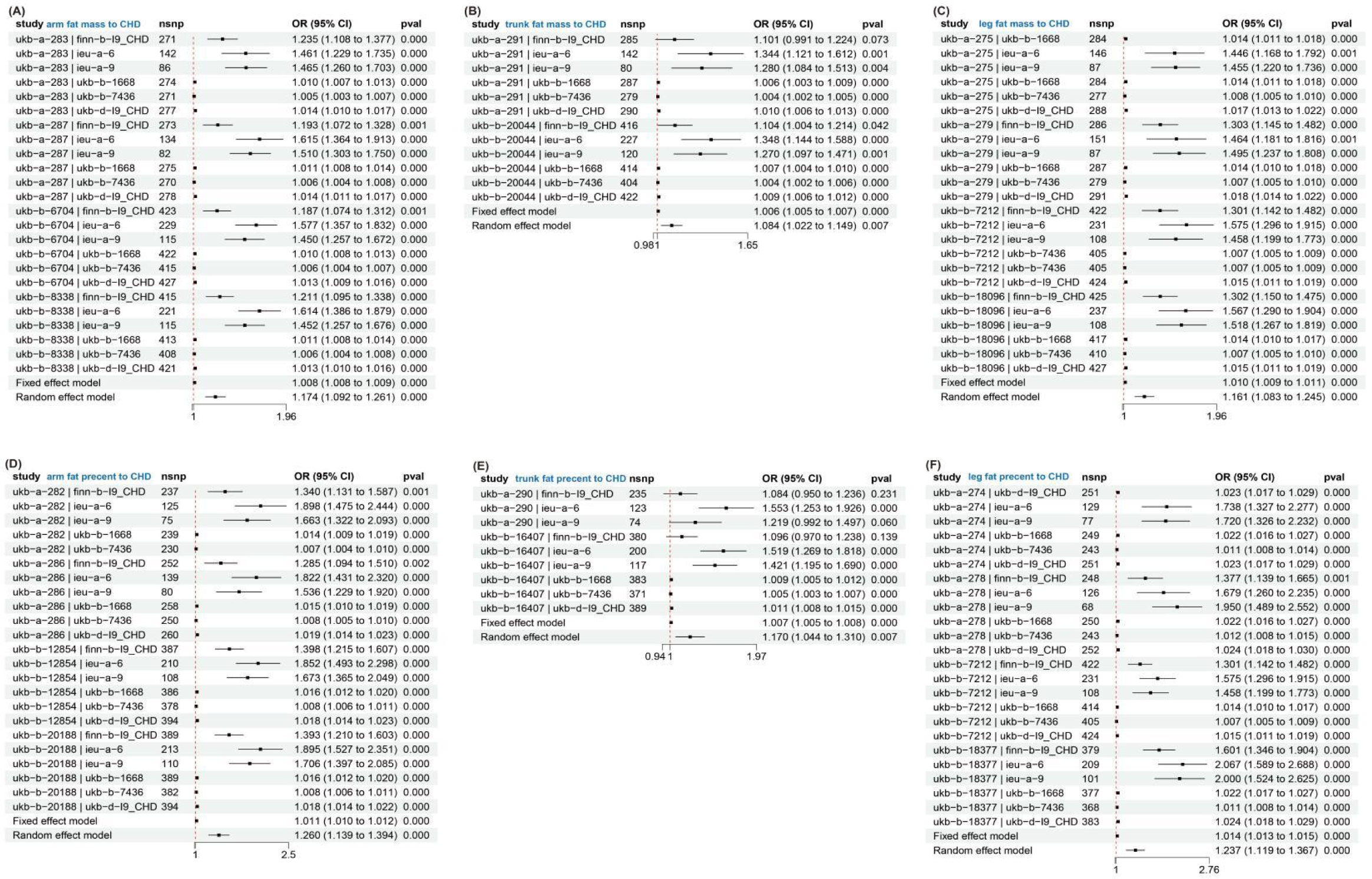
Mendelian randomization meta-analysis of the relationship between regional body fat and coronary heart disease. CHD, coronary heart disease; OR, odds ratio.

As mentioned earlier, previous studies have generally considered TFM as a health risk factor, whereas LFM is a protective factor. These health risks include metabolic syndrome, insulin resistance, high blood pressure, and overall risk of inflammation, among others. However, there has been limited research analyzing CVD or heart disease as endpoints. Trunk fat accumulates primarily in the chest, abdomen, and back. Its composition and effects on health are complex, and encompass the influence of both visceral and subcutaneous fat. However, the underlying mechanisms are not well understood. Current research suggests that visceral fat in the trunk region secretes more free fatty acids^29,30^ but has reduced adiponectin expression and secretion rates^31–34^, leading to insulin resistance^35,36^, concentric left ventricular remodeling^37^, and left atrial dysfunction^38^. Deep subcutaneous fat in the trunk exhibits a phenotype similar to that of visceral fat, whereas superficial subcutaneous fat in the legs possesses stronger metabolic characteristics^39–41^. The results of this study are consistent with those of previous studies showing a significant association between increased TFM and both cardiovascular risk factors and CHD **(TableS3)**. After logistic regression, subgroup analysis, and MR meta-analysis, the performance of the TFM remained consistently stable, with a weakened association with the outcomes observed only in two subgroups: PIR<1 and high blood pressure. Although TFP also demonstrated a causal relationship in the MR meta-analysis, it showed moderate performance in logistic regression analysis and had limited clinical significance **(Table3).** Thus, we believe that TFM is a risk factor for heart disease and has a clinical value in risk screening.

Previous studies have commonly regarded leg fat as a protective factor against cardiovascular risk; however, this study did not find an association between leg fat and CVD or heart disease. Indeed, RCS and MR meta-analyses suggested that leg fat is a risk factor for heart disease. First, from the baseline survey, it is evident that different RFMs in the body are positively correlated. With an increase in the LFM, the other RFMs also increase (**TableS1-TableS4**). The Pearson correlation coefficient revealed that LFM was strongly correlated with AFM and TFM, but weakly correlated with HFM **(Table2)**. The protective effect of leg fat may be overshadowed by risk effects in other regions. This could also explain why the results of the RCS and MR meta-analyses indicate that LFM is a risk factor for heart disease. Correcting for other RFMs before analyzing the relationship between LFM and heart disease was not the aim of this study, and it has no practical significance for the risk assessment of heart disease. Second, previous studies did not focus on the disease itself as the endpoint but rather on risk factors such as dyslipidemia^26,42^, insulin resistance^43–45^, cardiometabolic functions^19,46,47^, and inflammatory markers^16^. Leg fat itself may not be a suitable risk indicator for CVD; it merely affects the upstream indicators of CVD, which was the aim of this study. Third, most previous studies focused on the ratio of trunk to leg fat, not on leg fat alone ^19,20,42,44,46,47^. This study was an independent investigation of fat in various regions of the body. Fourth, although leg fat has been shown to be beneficial for some aspects of cardiovascular health, the population with high LFM in this study was more prone to obesity, hypertension, and even diabetes **(TableS4)**. However, whether these risk factors are associated with LFM remains unclear. This could also explain why LFM appeared to be a risk factor for heart disease in the RCS and MR meta-analyses as these two analyses did not adequately adjust for confounding factors. Finally, the study population consisted of middle-aged and young adults aged 20–59 years, which differed from the populations studied in previous studies. With advancing age, there are changes in fat distribution patterns. Older adults often experience a loss of fat in their limbs and an increase in abdominal fat. Therefore, leg fat may be more important in older individuals than in middle-aged or young adults. In conclusion, we believe that LFM has no value in risk screening for heart disease, and further research is needed to explore its relationship with cardiovascular health.

This study utilized nationally representative data and an MR meta-analysis, making it the first comprehensive analysis of the relationship between regional body fat and heart disease. This is the first study to target middle-aged and young adults. Overall, based on the results of logistic regression, HFM, HFP, AFM, and TFM were all correlated with heart disease, with only a slight decrease in correlation in some subgroups. Currently, there is very little research on head fat, and in our study, it was a highly promising indicator. Specifically, for every 0.1 kg increase in HFM, the risk of heart disease increased by 31%, whereas for every 1% increase in HFP, the risk of heart disease increased by 37% **(Table3)**. HFP exhibited even more stable performance in the subgroup analysis. In Pearson correlation analysis, the correlation between head fat and fat in other regions was not strong **(Table2)**, indicating that the influence of head fat on heart disease was relatively independent of other regions. Unfortunately, there is no information on HFM and HFP in the IEU OpenGWAS project, leading to a lack of MR meta-analyses specifically for head fat. However, by combining MR analysis with other RFMs, we believe there is a strong likelihood of a causal relationship between head fat and heart disease. The primary aim of this study was to establish the correlation between regional body fat and cardiovascular diseases in middle-aged and young adults, with the goal of providing a basis for the early screening of cardiovascular diseases in this population. Measurement of regional fat is convenient, economical, and low in radiation, making it suitable for community-based settings.

This study had certain limitations. First, in conducting the correlation analysis, we did not consider the mutual influence between different RFMs because this was not the primary focus of this study. This study aimed to analyze the relationship between body RFMs and heart disease to understand which RFM would be suitable as screening indicators for heart disease in the real world. Second, the MR analysis in this study was not solely focused on middle-aged and young adults; hence, the population characteristics did not fully match those of observational studies. Third, when conducting an MR analysis, it is generally preferred to have different populations for exposure and outcomes. However, due to the scarcity of relevant studies, we included all studies from the IEU OpenGWAS for analysis. In this study, the MR analysis, serving as a supplementary analysis to observational studies, could not conclusively prove causality but offered substantial potential.

## CONCLUSIONS

In conclusion, we believe that HFM, HFP, AFM, and TFM are risk factors for the onset of heart disease in middle-aged and young adults, and that there may be a causal relationship. Our study may provide new references for the early screening and intervention of heart disease in the middle-aged and young population.

## Data Availability

Interested researchers may contact the corresponding author via email to request access to the data. Please specify the intended use of the data and any necessary qualifications in your email. We will share the data as much as possible, based on reasonable requests and in accordance with data sharing agreements.

## ACKNOWLEDGMENTS

None

## SOURCES OF FUNDING

The funding for this study was provided by the Central Guiding Local Science and Technology Development Fund of Liaoning Province, China. The project is titled “Construction Research on Public Cardiovascular and Cerebrovascular Emergency Sudden Death Prevention and Treatment System Based on Intelligent Emergency Platform.” Project Number: 2023JH6/100200001.

## DISCLOSURES

None

## Non-standard Abbreviations and Acronyms

NHANES: National Health and Nutrition Examination Survey
CVD: cardiovascular disease
CHD: coronary heart disease
MR: mendelian randomization
SNPs: Single Nucleotide Polymorphisms
DXA: Dual-Energy X-ray Absorptiometry
RFM: regional fat mass
HFM: head fat mass
AFM: arm fat mass
TFM: trunk fat mass
LFM: leg fat mass
RFP: regional fat percentage
HFP: head fat percentage
AFP: arm fat percentage
TFP: trunk fat percentage
LFP: leg fat percentage
RCS: restricted cubic splines
MCQ: medical conditions questionnaire
BMI: body mass index
IVW: inverse variance-weighted
PIR: poverty income ratio
OR: odds ratio

## Notes

### Competing Interest Statement

The authors have declared no competing interest.

### Clinical Trial

This study is a cross-sectional study based on public databases and does not require clinical registration.

### Author Declarations

The Medical Scientific Research Ethics Committee of the First Hospital of China Medical University

## REFERENCES

1. Arnett DK, Blumenthal RS, Albert MA, Buroker AB, Goldberger ZD, Hahn EJ, Himmelfarb CD, Khera A, Lloyd-Jones D, McEvoy JW, et al. 2019 ACC/AHA Guideline on the Primary Prevention of Cardiovascular Disease: A Report of the American College of Cardiology/American Heart Association Task Force on Clinical Practice Guidelines. Circulation. 2019;140:e596–e646. doi: 10.1161/cir.0000000000000678

2. Zhao D, Liu J, Wang M, Zhang X, Zhou M. Epidemiology of cardiovascular disease in China: current features and implications. Nature reviews Cardiology. 2019;16:203–212. doi: 10.1038/s41569-018-0119-4

3. Turk-Adawi K, Sarrafzadegan N, Fadhil I, Taubert K, Sadeghi M, Wenger NK, Tan NS, Grace SL. Cardiovascular disease in the Eastern Mediterranean region: epidemiology and risk factor burden. Nature reviews Cardiology. 2018;15:106–119. doi: 10.1038/nrcardio.2017.138

4. Piepoli MF, Hoes AW, Agewall S, Albus C, Brotons C, Catapano AL, Cooney MT, Corrà U, Cosyns B, Deaton C, et al. 2016 European Guidelines on cardiovascular disease prevention in clinical practice: The Sixth Joint Task Force of the European Society of Cardiology and Other Societies on Cardiovascular Disease Prevention in Clinical Practice (constituted by representatives of 10 societies and by invited experts)Developed with the special contribution of the European Association for Cardiovascular Prevention & Rehabilitation (EACPR). European heart journal. 2016;37:2315–2381. doi: 10.1093/eurheartj/ehw106

5. Sacco RL, Roth GA, Reddy KS, Arnett DK, Bonita R, Gaziano TA, Heidenreich PA, Huffman MD, Mayosi BM, Mendis S, et al. The Heart of 25 by 25: Achieving the Goal of Reducing Global and Regional Premature Deaths From Cardiovascular Diseases and Stroke: A Modeling Study From the American Heart Association and World Heart Federation. Circulation. 2016;133:e674–690. doi: 10.1161/cir.0000000000000395

6. Roth GA, Nguyen G, Forouzanfar MH, Mokdad AH, Naghavi M, Murray CJ. Estimates of global and regional premature cardiovascular mortality in 2025. Circulation. 2015;132:1270–1282. doi: 10.1161/circulationaha.115.016021

7. Després JP, Moorjani S, Lupien PJ, Tremblay A, Nadeau A, Bouchard C. Regional distribution of body fat, plasma lipoproteins, and cardiovascular disease. Arteriosclerosis (Dallas, Tex). 1990;10:497–511. doi: 10.1161/01.atv.10.4.497

8. Canoy D, Boekholdt SM, Wareham N, Luben R, Welch A, Bingham S, Buchan I, Day N, Khaw KT. Body fat distribution and risk of coronary heart disease in men and women in the European Prospective Investigation Into Cancer and Nutrition in Norfolk cohort: a population-based prospective study. Circulation. 2007;116:2933–2943. doi: 10.1161/circulationaha.106.673756

9. Lim S, Meigs JB. Ectopic fat and cardiometabolic and vascular risk. International journal of cardiology. 2013;169:166–176. doi: 10.1016/j.ijcard.2013.08.077

10. Wajchenberg BL. Subcutaneous and visceral adipose tissue: their relation to the metabolic syndrome. Endocrine reviews. 2000;21:697–738. doi: 10.1210/edrv.21.6.0415

11. Després JP, Lemieux I. Abdominal obesity and metabolic syndrome. Nature. 2006;444:881–887. doi: 10.1038/nature05488

12. Zhang X, Hu EA, Wu H, Malik V, Sun Q. Associations of leg fat accumulation with adiposity-related biological factors and risk of metabolic syndrome. Obesity (Silver Spring, Md). 2013;21:824–830. doi: 10.1002/oby.20028

13. Carey VJ, Walters EE, Colditz GA, Solomon CG, Willett WC, Rosner BA, Speizer FE, Manson JE. Body fat distribution and risk of non-insulin-dependent diabetes mellitus in women. The Nurses’ Health Study. American journal of epidemiology. 1997;145:614–619. doi: 10.1093/oxfordjournals.aje.a009158

14. Yang L, Deng H, Pan W, Huang X, Xu K, Zhang X, Hu X, Gu X. The Inverse Association of Leg Fat Mass and Osteoporosis in Individuals with Type 2 Diabetes Independent of Lean Mass. Diabetes, metabolic syndrome and obesity: targets and therapy. 2022;15:1321–1330. doi: 10.2147/dmso.S358717

15. Hayashi T, Boyko EJ, Leonetti DL, McNeely MJ, Newell-Morris L, Kahn SE, Fujimoto WY. Visceral adiposity and the prevalence of hypertension in Japanese Americans. Circulation. 2003;108:1718–1723. doi: 10.1161/01.Cir.0000087597.59169.8d

16. Wu H, Qi Q, Yu Z, Sun Q, Wang J, Franco OH, Sun L, Li H, Liu Y, Hu FB, et al. Independent and opposite associations of trunk and leg fat depots with adipokines inflammatory markers, and metabolic syndrome in middle-aged and older Chinese men and women. The Journal of clinical endocrinology and metabolism. 2010;95:4389–4398. doi: 10.1210/jc.2010-0181

17. Barbour-Tuck EN, Erlandson MC, Sherar LB, Eisenmann JC, Muhajarine N, Foulds H, Vatanparast H, Nisbet C, Kontulainen S, Baxter-Jones ADG. Relationship Between Trajectories of Trunk Fat Development in Emerging Adulthood and Cardiometabolic Risk at 36 Years of Age. Obesity (Silver Spring, Md). 2019;27:1652–1660. doi: 10.1002/oby.22576

18. He Q, Zhang X, He S, Gong L, Sun Y, Heshka S, Deckelbaum RJ, Gallagher D. Higher insulin, triglycerides, and blood pressure with greater trunk fat in Tanner 1 Chinese. Obesity (Silver Spring, Md). 2007;15:1004–1011. doi: 10.1038/oby.2007.599

19. Cioffi CE, Alvarez JA, Welsh JA, Vos MB. Truncal-to-leg fat ratio and cardiometabolic disease risk factors in US adolescents: NHANES 2003-2006. Pediatric obesity. 2019;14:e12509. doi: 10.1111/ijpo.12509

20. Han E, Cho NH, Kim MK, Kim HS. Lower Leg Fat Depots Are Associated with Albuminuria Independently of Obesity, Insulin Resistance, and Metabolic Syndrome (Korea National Health and Nutrition Examination Surveys 2008 to 2011). Diabetes & metabolism journal. 2019;43:461–473. doi: 10.4093/dmj.2018.0081

21. Saunders TJ, Davidson LE, Janiszewski PM, Després JP, Hudson R, Ross R. Associations of the limb fat to trunk fat ratio with markers of cardiometabolic risk in elderly men and women. The journals of gerontology Series A, Biological sciences and medical sciences. 2009;64:1066–1070. doi: 10.1093/gerona/glp079

22. Kim JH, Chon J, Soh Y, Han YR, Won CW, Lee SA. Trunk fat mass correlates with balance and physical performance in a community-dwelling elderly population: Results from the Korean Frailty and aging cohort study. Medicine (Baltimore). 2020;99:e19245. doi: 10.1097/md.0000000000019245

23. Smith GD, Ebrahim S. ’Mendelian randomization’: can genetic epidemiology contribute to understanding environmental determinants of disease? International journal of epidemiology. 2003;32:1–22. doi: 10.1093/ije/dyg070

24. Smith GD, Timpson N, Ebrahim S. Strengthening causal inference in cardiovascular epidemiology through Mendelian randomization. Annals of medicine. 2008;40:524–541. doi: 10.1080/07853890802010709

25. Wang XC, Liu H, Huang YY, Sun H, Bu L, Qu S. Head fat is a novel method of measuring metabolic disorder in Chinese obese patients. Lipids in health and disease. 2014;13:113. doi: 10.1186/1476-511x-13-113

26. Sánchez-López M, Ortega FB, Moya-Martínez P, López-Martínez S, Ortiz-Galeano I, Gómez-Marcos MA, Sjöström M, Martínez-Vizcaíno V. Leg fat might be more protective than arm fat in relation to lipid profile. European journal of nutrition. 2013;52:489–495. doi: 10.1007/s00394-012-0350-4

27. Shi J, Yang Z, Niu Y, Zhang W, Li X, Zhang H, Lin N, Gu H, Wen J, Ning G, et al. Large mid-upper arm circumference is associated with metabolic syndrome in middle-aged and elderly individuals: a community-based study. BMC Endocr Disord. 2020;20:78. doi: 10.1186/s12902-020-00559-8

28. Gavi S, Feiner JJ, Melendez MM, Mynarcik DC, Gelato MC, McNurlan MA. Limb fat to trunk fat ratio in elderly persons is a strong determinant of insulin resistance and adiponectin levels. The journals of gerontology Series A, Biological sciences and medical sciences. 2007;62:997–1001. doi: 10.1093/gerona/62.9.997

29. Arner P. Insulin resistance in type 2 diabetes: role of fatty acids. Diabetes/metabilosm research and reviews. 2002;18 Suppl 2:S5–9. doi: 10.1002/dmrr.254

30. Kissebah AH, Krakower GR. Regional adiposity and morbidity. Physiological reviews. 1994;74:761–811. doi: 10.1152/physrev.1994.74.4.761

31. Manolopoulos KN, Karpe F, Frayn KN. Gluteofemoral body fat as a determinant of metabolic health. International journal of obesity (2005). 2010;34:949–959. doi: 10.1038/ijo.2009.286

32. Motoshima H, Wu X, Sinha MK, Hardy VE, Rosato EL, Barbot DJ, Rosato FE, Goldstein BJ. Differential regulation of adiponectin secretion from cultured human omental and subcutaneous adipocytes: effects of insulin and rosiglitazone. The Journal of clinical endocrinology and metabolism. 2002;87:5662–5667. doi: 10.1210/jc.2002-020635

33. Lihn AS, Bruun JM, He G, Pedersen SB, Jensen PF, Richelsen B. Lower expression of adiponectin mRNA in visceral adipose tissue in lean and obese subjects. Molecular and cellular endocrinology. 2004;219:9–15. doi: 10.1016/j.mce.2004.03.002

34. Després JP. Is visceral obesity the cause of the metabolic syndrome? Annals of medicine. 2006;38:52–63. doi: 10.1080/07853890500383895

35. Cnop M, Landchild MJ, Vidal J, Havel PJ, Knowles NG, Carr DR, Wang F, Hull RL, Boyko EJ, Retzlaff BM, et al. The concurrent accumulation of intra-abdominal and subcutaneous fat explains the association between insulin resistance and plasma leptin concentrations: distinct metabolic effects of two fat compartments. Diabetes. 2002;51:1005–1015. doi: 10.2337/diabetes.51.4.1005

36. Kim G, Jo K, Kim KJ, Lee YH, Han E, Yoon HJ, Wang HJ, Kang ES, Yun M. Visceral adiposity is associated with altered myocardial glucose uptake measured by (18)FDG-PET in 346 subjects with normal glucose tolerance, prediabetes, and type 2 diabetes. Cardiovascular diabetology. 2015;14:148. doi: 10.1186/s12933-015-0310-4

37. Neeland IJ, Gupta S, Ayers CR, Turer AT, Rame JE, Das SR, Berry JD, Khera A, McGuire DK, Vega GL, et al. Relation of regional fat distribution to left ventricular structure and function. Circulation Cardiovascular imaging. 2013;6:800–807. doi: 10.1161/circimaging.113.000532

38. Evin M, Broadhouse KM, Callaghan FM, McGrath RT, Glastras S, Kozor R, Hocking SL, Lamy J, Redheuil A, Kachenoura N, et al. Impact of obesity and epicardial fat on early left atrial dysfunction assessed by cardiac MRI strain analysis. Cardiovascular diabetology. 2016;15:164. doi: 10.1186/s12933-016-0481-7

39. Marinou K, Hodson L, Vasan SK, Fielding BA, Banerjee R, Brismar K, Koutsilieris M, Clark A, Neville MJ, Karpe F. Structural and functional properties of deep abdominal subcutaneous adipose tissue explain its association with insulin resistance and cardiovascular risk in men. Diabetes care. 2014;37:821–829. doi: 10.2337/dc13-1353

40. Peppa M, Koliaki C, Hadjidakis DI, Garoflos E, Papaefstathiou A, Katsilambros N, Raptis SA, Dimitriadis GD. Regional fat distribution and cardiometabolic risk in healthy postmenopausal women. Eur J Intern Med. 2013;24:824–831. doi: 10.1016/j.ejim.2013.07.001

41. Goss AM, Gower BA. Insulin sensitivity is associated with thigh adipose tissue distribution in healthy postmenopausal women. Metabolism: clinical and experimental. 2012;61:1817–1823. doi: 10.1016/j.metabol.2012.05.016

42. Duran I, Martakis K, Alberg E, Jackels M, Ewert KR, Schoenau E. Association of Trunk/Leg Fat Mass Ratio with Low-Density Lipoproteins-Cholesterol and Triglycerides Concentration in Children and Adolescents: A Cross-Sectional, Retrospective Study. Childhood obesity (Print). 2020;16:428–439. doi: 10.1089/chi.2019.0307

43. Samouda H, De Beaufort C, Stranges S, Hirsch M, Van Nieuwenhuyse JP, Dooms G, Gilson G, Keunen O, Leite S, Vaillant M, et al. Cardiometabolic risk: leg fat is protective during childhood. Pediatric diabetes. 2016;17:300–308. doi: 10.1111/pedi.12292

44. Cui B, Li W, Wang G, Li P, Zhu L, Zhu S. The predictive value of trunk/leg fat ratio for type 2 diabetes mellitus remission after bariatric surgery: A new observation and insight. Front Endocrinol (Lausanne). 2022;13:1068917. doi: 10.3389/fendo.2022.1068917

45. Snijder MB, Dekker JM, Visser M, Bouter LM, Stehouwer CD, Yudkin JS, Heine RJ, Nijpels G, Seidell JC. Trunk fat and leg fat have independent and opposite associations with fasting and postload glucose levels: the Hoorn study. Diabetes care. 2004;27:372–377. doi: 10.2337/diacare.27.2.372

46. Han E, Lee YH, Lee BW, Kang ES, Lee IK, Cha BS. Anatomic fat depots and cardiovascular risk: a focus on the leg fat using nationwide surveys (KNHANES 2008-2011). Cardiovascular diabetology. 2017;16:54. doi: 10.1186/s12933-017-0536-4

47. Kishimoto I. Trunk-to-Leg Fat Ratio - An Emerging Early Marker of Childhood Adiposity, and Future Cardiometabolic Risks. Circulation journal: official journal of the Japanese Circulation Society. 2016;80:1707–1709. doi: 10.1253/circj.CJ-16-0635

